# HGGT: Heterogeneous Gated Graph Transformer for Predicting Clinical Trial Success

**DOI:** 10.64898/2026.06.28.26356795

**Authors:** Long Qian, Xin Lu, Parvez Haris, Yingjie Yang

## Abstract

Clinical trials are critical milestones in the drug development pipeline, yet their high failure rates and substantial costs underscore the need for robust predictive models. This study introduces a Heterogeneous Gated Graph Transformer (HGGT) model tailored to predict clinical trial success. Unlike existing methods that typically model trial-related entities in isolation or with homogeneous graphs, HGGT explicitly models the rich heterogeneous relationships among trials, diseases, drugs, genes, targets, abstracts, and eligibility criteria through a gated graph transformer architecture, which dynamically learns and weights multi-type relational interactions to capture complex biological and clinical dependencies. By integrating heterogeneous graph representation with transformer-based context modeling, HGGT effectively captures non-linear, multi-scale interactions across biomedical entities, leading to improved predictive performance for trial success. Experimental results demonstrate that the HGGT model achieves strong performance, with the highest PR-AUC, F1 score, and ROC-AUC across three phases. These findings highlight the potential of graph-based deep learning approaches in optimizing clinical trial design and resource allocation, ultimately accelerating the translation of novel therapies into clinical practice.

## 1. Introduction

The drug development process is a lengthy, costly, and high-risk endeavor, with clinical trials representing a critical bottleneck. It is estimated that only 10% of candidate drugs advance from preclinical testing to regulatory approval, with failure rates varying across trial phases. Specifically, Phase I trials focus on safety and dosage, Phase II on efficacy and side effects, and Phase III on confirming effectiveness in larger populations. Accurate prediction of trial success at each phase could significantly reduce costs, estimated at over $2 billion per approved drug, and streamline the development timeline [1]. Identifying reliable predictors of trial outcome is therefore paramount, and genetic evidence emerges as a key determinant in this regard. The findings show that trials lacking strong genetic evidence are more likely to be terminated for lack of efficacy. Trials that are not supported by genetic data from human populations or genetically modified animal models are more likely to fail. In addition, trials are more likely to be stopped for safety reasons if the drug target gene is highly restricted in the human population or widely expressed in tissues [2].

Earlier attempts were often made to predict individual components like antidepressants in clinical trials to improve the trial results for each trial [3]. Recently, researchers started to propose general methods for trial outcome predictions. For example, Lo et al. [4] predicted drug approvals for 15 disease groups based on drug and clinical trial features using classical machine learning methods. SPOT [5] employed a meta-learning technique to organize trials with the same subject into a chronological sequence, leveraging insights from related trials and their predictive advancements. However, traditional predictive models for clinical trial outcomes have relied on the approaches that often overlook the complex interdependencies between biological entities (e.g., drugs, diseases, genes) and trial-specific entities (e.g., inclusion criteria, study design).

Graph neural networks have emerged as powerful tools for modeling relational data, with heterogeneous graph models capable of handling multiple node and edge types—an essential feature given the diverse entities involved in clinical trials [3]. In heterogeneous graphs, a range of techniques have been developed. HAN [6] applies attention mechanisms at both the node and semantic levels to evaluate the importance of individual nodes and their neighbors connected via meta-paths, as well as the significance of various meta-paths. HetGNN [7] adopts a method to model both structural and content-based heterogeneity. It uses random walk sampling to select closely related heterogeneous neighbors, which are then categorized according to node type. These sampled nodes are processed by a two-component neural network designed to incorporate both structural and content differences, resulting in node embeddings. MHGCN [8] is designed to automatically detect meaningful heterogeneous interactions along meta-paths of different lengths in multiplex heterogeneous networks through multi-layer convolutional aggregation, producing node representations that combine structural and semantic insights. However, MHGCN faces limitations in terms of scalability. HGT [9] differentiates itself by modeling heterogeneous nodes and edges using meta-relations, with parameters tailored to specific node and edge types. This method allows the discovery of shared and distinctive patterns across diverse relationships without relying on pre-specified meta-paths, thereby avoiding the need to determine the most relevant meta-paths in biological settings.

Recent successes in graph neural network-based approaches have spurred interest in utilizing biological networks to exploit complementary biology data. For example, The layer attention graph convolutional network (LAGCN) method, as proposed by Yu et al. [10], establishes a drug–drug similarity network by applying the Jaccard index to various drug-related datasets, including targets, pathways, and substructures, all of which are sourced from the DrugBank database [11]. Peng et al. [12] emphasized the necessity of considering gene-to-gene link prediction and thus developed the MTGCN multi-task framework which is a multi-task learning approach based on graph convolutional neural networks to predict cancer driver genes. A most related work using GNN in clinical trial outcome prediction is HINT [13]: which is graph neural network for feature interactions. Then, it feeds the interacted embeddings to a prediction model for outcome predictions.

Our work builds on these advancements while addressing key limitations in existing approaches for clinical trial success prediction. Although conventional graph neural networks (GNNs) have been applied to model biomedical relationships, most either assume homogeneous graph structures or use fixed, non-adaptive relation weighting, which fails to account for the diverse and context-dependent importance of different entity and relation types in clinical trials. In response, we develop a trial-tailored Heterogeneous Gated Graph Transformer (HGGT) model that explicitly captures the unique relationships between trials and their associated biological and clinical entities. Unlike standard GNNs, HGGT incorporates gated attention mechanisms that adaptively weight node and edge types according to their clinical and biological relevance, rather than applying uniform or predefined relation weights. This adaptive weighting enables more nuanced and context-aware modeling of the complex systems underlying clinical trials, making our approach better suited for capturing the heterogeneous, multi-scale interactions that determine trial success.

Also, current clinical datasets lack genetic information, which may lead to an incomplete understanding of disease mechanisms and limited accuracy in clinical prediction. TRIALPANORAMA [14] is a structured and extensive database and benchmark designed to support AI-driven activities in clinical trial processes, including systematic reviews and the design of trials. It integrates normalized elements such as trial setups, treatments, diseases, biomarkers, and efficacy endpoints, along with linkages to well-known biomedical ontologies like DrugBank and MedDRA. We extend the TOP dataset [13], a clinical trial outcome prediction benchmark, by integrating an additional clinical knowledge graph sourced from the TRIALPANORAMA database. This augmented dataset, named TOP-gene, incorporates target, gene, and abstract information for specific clinical trials.

In this study, our contributions are threefold:

1. a new combined dataset termed TOP-gene is developed to enable clinical trial outcome prediction leveraging genetic information. Specifically, we integrated external databases to enrich the original TOP dataset with comprehensive gene-related and target-specific information.
2. A novel Heterogeneous Gated Graph Transformer is proposed for clinical trial outcome prediction. Unlike existing heterogeneous graph models that apply uniform non-adaptive relation weighting and overlook feature space differences across biomedical entities, this model is tailored with trial-centric gated feature filtering and type-specific feature projection. It captures dedicated representations for different node and edge types, effectively addressing the inherent heterogeneity of the constructed clinical trial graph.
3. We evaluated HGGT achieved the best PR-AUC, F1, ROC-AUC scores on phase I-. II-, and III-level predictions, respectively demonstrating the model’s predictive performance against state-of-the-art baselines using real-world data. Our work establishes that genetic factors can serve as an effective means to improve the performance of AI models.

## 3. Methods

### 3.1 Data processing

We obtained clinical trial data from TOP, comprising information on drugs, diseases, and eligibility criteria across a total of 12,465 trials. These trials were categorized into three phases: Phase I (1,787 trials), Phase II (6,102 trials), and Phase III (4,576 trials). Subsequently, we integrated gene, target, and abstract information using the drug_moa table from the TRIALPANORAMA database [14], based on matching drug identifiers. Mechanism of action (MoA) information is represented through the drug_moa_id field, which links each drug to its corresponding biological targets—such as enzymes, receptors, or transporters—along with their interaction types (e.g., inhibitor, agonist).

### 3.2 Graph Construction

To effectively capture the domain-specific knowledge in clinical trial data, we extend node and edge types with fine-grained biomedical semantics. We customize 6 clinical trial-oriented node types including trial, criteria, SMILES, ICD code, gene and target which map directly to core biomedical trial entities for precise domain knowledge representation. Additionally, we design 9 domain-specific semantic edges such as Gene encodes target and Trial has disease that encode relational characteristics between these nodes, enhancing the model’s ability to capture biomedical semantic correlations as shown in Figure 1.

**Figure 1.**
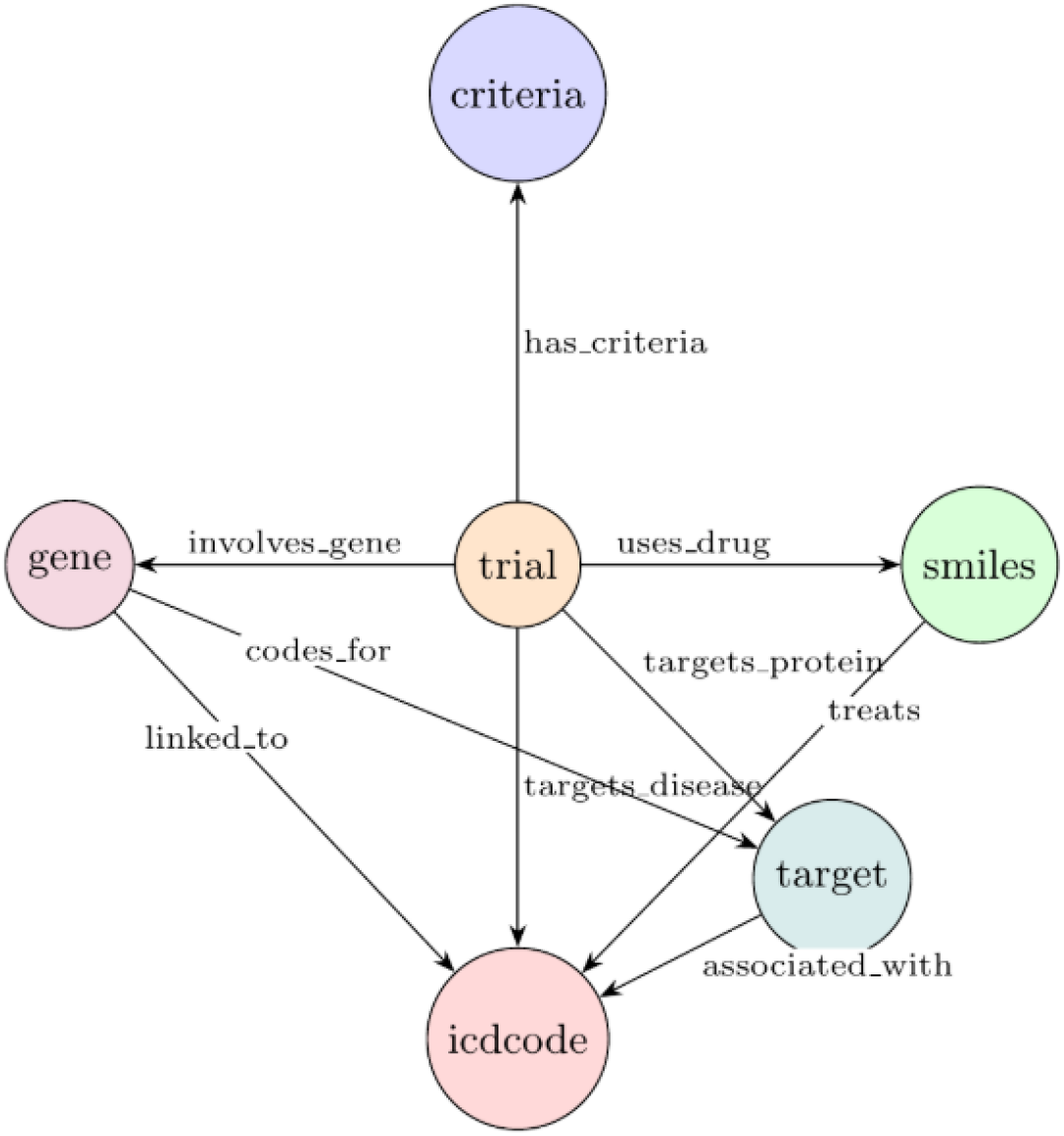
Heterogeneous graph structures in capturing domain-specific knowledge of clinical trials. Criteria are the inclusion/exclusion criteria text, and ICD code is the disease classification.

### 3.3 Model Architecture

#### 3.3.1 Input Layers

We use specific encoder of each modal for the initial embedding of HGGT. Criteria and abstract features were generated using BioBERT [15], a pre-trained language model fine-tuned on biomedical text, to capture semantic information from entity names and text (e.g., abstracts, criteria). Long text features were processed by segmenting and averaging embeddings to handle length constraints. It processes both inclusion and exclusion criteria separately, generating 768-dimensional criteria embedding for each, which are subsequently concatenated.

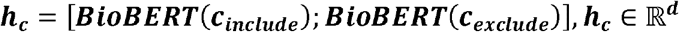

For disease representation, we implement a Graph-based Attention Model (GRAM) [16] that captures the hierarchical nature of ICD-10 codes (disease code), producing a mean-pooling of all code embeddings.

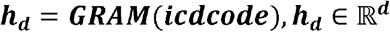

Drug compounds (represented by SMILES strings) in the treatment set are encoded using a graph message passing neural network (MPNN) [17], followed by mean-pooling to obtain the treatment embedding

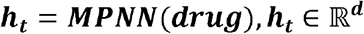

The original Heterogeneous Graph Transformer (HGT) model presupposes that all node features possess identical input dimensionalities and employs only a simple linear transformation followed by a tanh activation function for type-specific feature adaptation shown in Figure 2. This assumption becomes unrealistic in our setting, where trial, criteria, molecular (SMILES), diagnosis (ICD code), gene and target nodes have fundamentally different feature spaces (e.g., 768⍰dim BioBERT text embeddings, 50⍰dim MPNN molecular embeddings, 128⍰dim GRAM hierarchical codes). To unify representations across diverse node types, we first design type⍰specific projection blocks. Each block takes raw features of a specific node type with input dimensions matched to the original feature size and maps them into a common shared latent space.

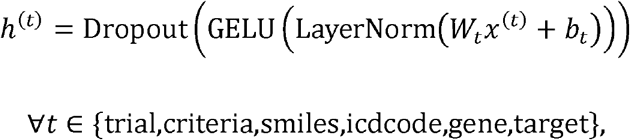

**Figure 2.**
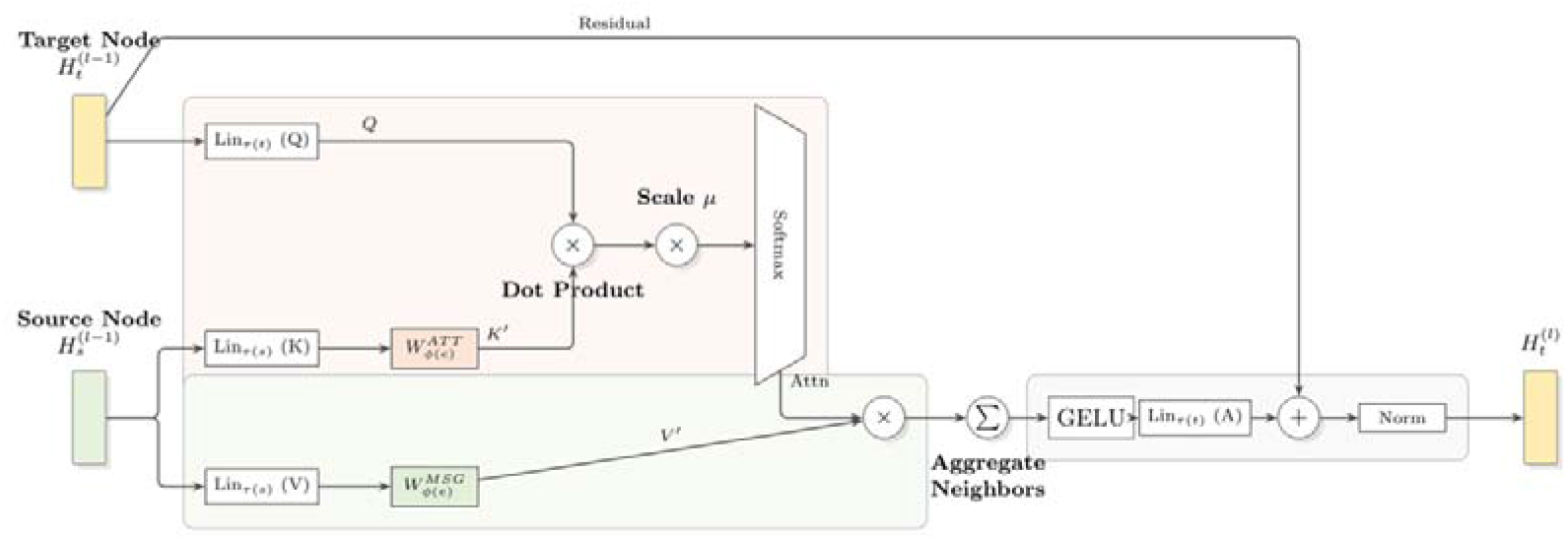
The standard heterogeneous graph Transformer layer is first used to encode the trial nodes and their heterogeneous neighbors, resulting in the experimental representation and representations of various neighbor types.

**Figure 3.**
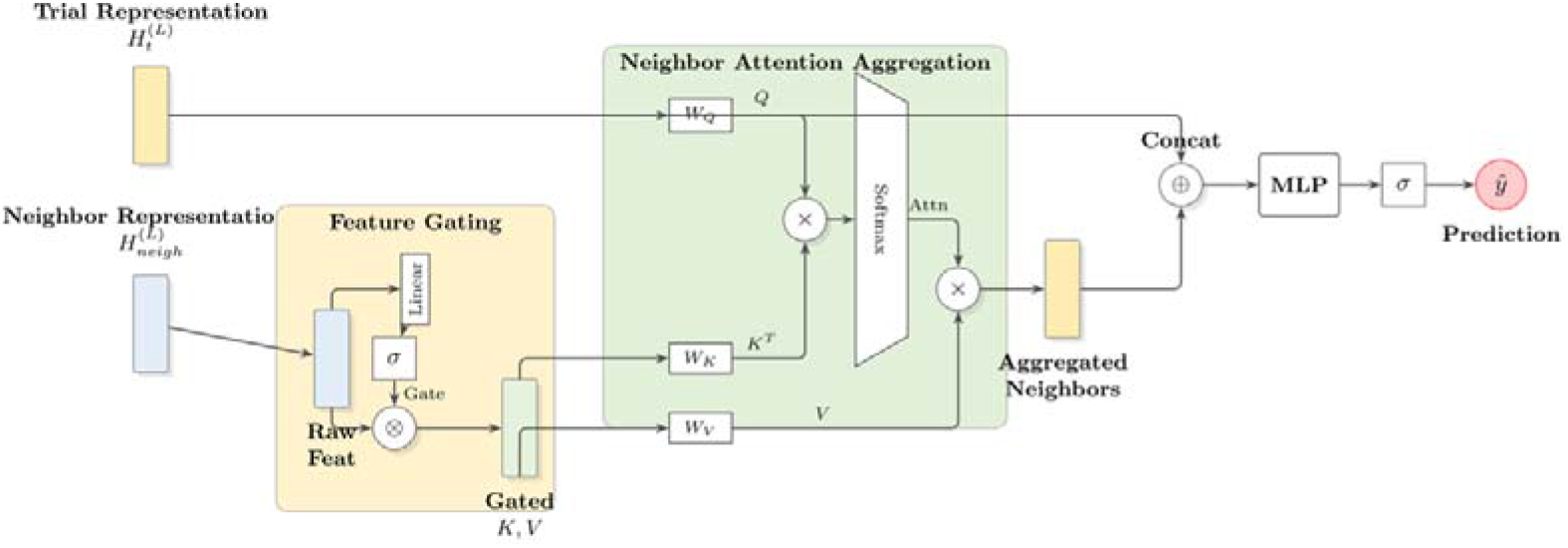
Architecture of the proposed feature gating and neighbor attention module for clinical trial outcome prediction. Neighbor representations (e.g., genes, targets) first undergo feature gating to weight and filter heterogeneous features. The trial node representation then acts as the query to perform attention aggregation on gated neighbors, adaptively emphasizing high-value information. Finally, the concatenated trial and aggregated neighbor representations are fed into a multi-layer perceptron to generate the prediction.

The architecture of each block is structured as a sequence of Linear, LayerNorm, GELU, and Dropout operations. This design offers enhanced non-linear processing and regularization capabilities compared to the conventional approach which utilizes a Linear layer followed by a tanh activation function.

#### 3.3.2 feature transformation

HGGT achieves differentiated processing of different types of nodes and edges through heterogeneous mutual attention mechanisms and meta-relation modeling, as shown in Figure 2. The core process follows a three-step process of “attention calculation - message passing aggregation - update”, which is formulaically expressed as:

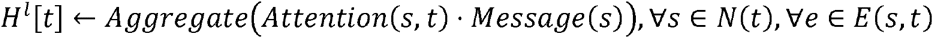

Instead of normal multi-head attention mechanism [18] [**Error! Reference source not found**.], we employ a Trial-Aware Attention Aggregation to dynamically fuse the gated neighbor’s representations. We treat the trial node’s own embedding *h*_*t*_ as the Query, and the gated neighbor features 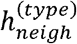 as both Keys and Values:

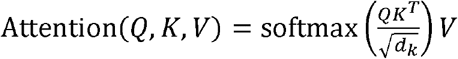

where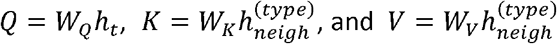

The learnable gating mechanism filters noise and emphasize informative features from heterogeneous neighbors. For a trial node *t* and its aggregated neighbor representation 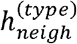 for a specific node type (e.g., Gene), we compute a gating score:

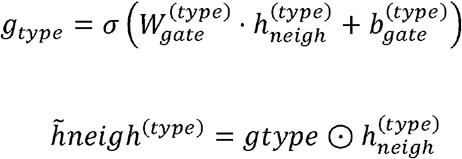

Where *σ* is the sigmoid function and ⊙ denotes element-wise multiplication. This allows the model to adaptively suppress irrelevant information (e.g., common but non-informative ICD codes) while amplifying critical signals.

Consequently, each layer not only computes standard message passing but also integrates this type-specific gating and relation-aware attention. This hierarchical aggregation allows the model to prioritize highly relevant relationships over less discriminative structural connections, significantly enhancing robustness against data noise and sparsity.

The new node features are

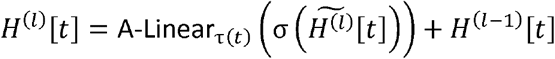

A-Linear_τ(*t*)_ is a linear transformation function applied to the aggregated feature vectors. σ is the activation function. *H*^(*l* −1)^ [*t*]is the node feature vector of the previous layer.

#### 3.3.3. Classification

The original HGT [9] adopts a shallow single-layer linear classifier with a log-softmax activation on top of node embeddings, which limits its ability to model complex clinical decision boundaries. In contrast, our model employs a deeper multi-layer MLP. This MLP takes the concatenation of a trial’s own embedding and its attention-aggregated neighbor representation as input and produces a fixed-dimensional feature vector. The classifier follows a 4*H* → 2*H* → *H*→ 1 progression, with LayerNorm, GELU and Dropout applied between intermediate layers. This design simultaneously enhances non-linear feature transformation and cross-type semantic fusion, while performing progressive dimensionality reduction with layer-wise regularization, thereby improving generalization without sacrificing discriminative power.

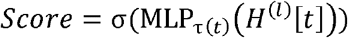

where MLP_τ(*t*)_is a transformation function applied to the aggregated feature vectors. σ is the sigmoid function. *H*^(*l*)^ [*t*]is the node feature vector of the previous layer. Score is the output of fully connected network.

#### 3.3.3 Loss Function

We train the model with a focal loss for binary classification. The binary focal loss over a mini⍰batch of size (N) is

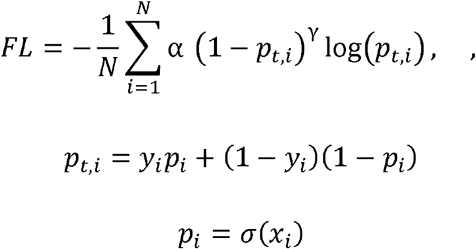

*p*_*t,i*_ is the predicted probability of the target class. *α* is a balancing factor used to adjust the importance of various elements. γ is the focusing parameter, which controls the degree of attention paid to difficult-to-classify samples.

### 3.4 Training Configuration

We employed the AdamW optimizer with an initial learning rate of 0.0001, weight decay (L2 regularization) of 0.01, beta coefficients (0.9, 0.999), and gradient clipping (max norm 1.0). The OneCycleLR scheduler was used for learning rate scheduling, featuring a maximum learning rate of, 10% warmup of total epochs, initial divisor 25.0, final divisor 1000.0, and cosine annealing. For label imbalance, BCEWithLogitsLoss with dynamic positive class weighting was adopted. Key hyperparameters included 1000 max epochs, batch size 32, early stopping (patience 30, improvement threshold), and random seed 42. Regularization strategies covered multi-level Dropout (0.2 for projection, 0.2–0.4 for classifiers), post-transformation Layer Normalization, gradient clipping, and weight decay.

## 4. Results

### 4.1 Baseline model

To demonstrate the superiority of HGGT, a comparison is made with the following methods:

- Logistic Regression (LR) [4]: A linear classification model implemented using scikit-learn with default hyperparameters.
- Random Forest (RF) [4]: An ensemble learning method based on decision trees, also implemented via scikit-learn with default settings.
- XGBoost [4]: An optimized implementation of gradient-boosted decision trees designed for computational efficiency and predictive performance.
- Adaptive Boosting (AdaBoost) [19]: A boosting-based ensemble method that iteratively adjusts sample weights to improve classification accuracy, implemented using scikit-learn.
- k-Nearest Neighbors + Random Forest (kNN+RF) [4]: A hybrid approach that employs kNN for missing data imputation followed by Random Forest for outcome prediction.
- Feedforward Neural Network (FFNN) [20]: A three-layer fully-connected neural network with hidden dimensions of 500 and 100, utilizing ReLU activation functions. The FFNN employs the same feature representation as HINT [13].
- Heterogeneous Graph Transformer (HGT) [9]: A graph neural network architecture specifically designed for modeling heterogeneous graphs with multiple node and edge types.
- DeepEnroll [21]: Originally developed for patient-trial matching, this model comprises three components: (1) a pre-trained BERT encoder for eligibility criteria embedding, (2) a hierarchical embedding module for disease information, and (3) an alignment mechanism to capture interactions between eligibility criteria and diseases. For trial outcome prediction, molecule embeddings generated by Message Passing Neural Networks (MPNN) are concatenated with the alignment model outputs.
- COMPOSE [22]: Similar to DeepEnroll, COMPOSE was initially proposed for patient-trial matching. It employs a convolutional neural network and a memory network to encode eligibility criteria and disease information, respectively. MPNN-derived molecule embeddings are similarly concatenated for trial outcome prediction.
- HINT [13]: An integrative framework incorporating multiple encoders: (1) an MPNN-based drug molecule encoder, (2) a GRAM-based disease ontology encoder, (3) a BERT-based trial eligibility criteria encoder, and (4) a pharmacokinetic encoder with graph neural networks for capturing feature interactions. The encoded representations are subsequently fed into a prediction module for outcome estimation.
- SPOT [5]: A meta-learning approach consisting of three stages: (1) identification of trial topics to cluster heterogeneous trial data from multiple sources, (2) generation and organization of trial embeddings by topic and timestamp to construct structured clinical trial sequences, and (3) treatment of each trial sequence as an independent task, enabling adaptation to new tasks with minimal parameter updates.

### 4.2 Model Performance

The HGGT model demonstrated strong predictive performance on TOP dataset as shown in Table 1, Table 2,and Table 3. HGT_aug denotes the original HGT model augmented with gene-level information, while HGGT_aug denotes our proposed HGGT architecture equipped with the same gene-level features.

**Table 1:** Performance comparison of different models on Phase I trials.

| Method | Phase I PR-AUC ± SD | Phase I F1 ± SD | Phase I ROC-AUC ± SD |
| --- | --- | --- | --- |
| LR | 0.500 ± 0.005 | 0.604 ± 0.005 | 0.520 ± 0.006 |
| RF | $0.518 \pm 0.005$ | $0.621 \pm 0.005$ | $0.525 \pm 0.006$ |
| XGBoost | $0.513 \pm 0.006$ | $0.621 \pm 0.007$ | $0.518 \pm 0.006$ |
| AdaBoost | $0.519 \pm 0.005$ | $0.622 \pm 0.007$ | $0.526 \pm 0.006$ |
| kNN + RF | $0.531 \pm 0.006$ | $0.625 \pm 0.007$ | $0.538 \pm 0.005$ |
| FFNN | $0.547 \pm 0.010$ | $0.634 \pm 0.015$ | $0.550 \pm 0.010$ |
| DeepEnroll | $0.568 \pm 0.007$ | $0.648 \pm 0.011$ | $0.575 \pm 0.013$ |
| COMPOSE | $0.564 \pm 0.007$ | $0.658 \pm 0.009$ | $0.571 \pm 0.011$ |
| HINT | $0.567 \pm 0.010$ | $0.665 \pm 0.010$ | $0.576 \pm 0.008$ |
| SPOT | $0.689 \pm 0.009$ | $0.714 \pm 0.011$ | $0.660 \pm 0.008$ |
| HGGT | $0.6191 \pm 0.0333$ | $0.6900 \pm 0.0154$ | $0.5615 \pm 0.0236$ |
| HGT_aug | $0.6742 \pm 0.0309$ | $0.6823 \pm 0.0163$ | $0.6256 \pm 0.0259$ |
| HGGT_aug | $0.6849 \pm 0.0305$ | $0.7126 \pm 0.0187$ | $0.6425 \pm 0.0204$ |

**Table 2:** Performance comparison of different models on Phase II trials.

| Method | Phase II PR-AUC ± SD | Phase II F1 ± SD | Phase II ROC-AUC ± SD |
| --- | --- | --- | --- |
| LR | $0.565 \pm 0.005$ | $0.555 \pm 0.006$ | $0.587 \pm 0.009$ |
| RF | $0.578 \pm 0.008$ | $0.563 \pm 0.009$ | $0.588 \pm 0.009$ |
| XGBoost | $0.586 \pm 0.006$ | $0.570 \pm 0.009$ | $0.600 \pm 0.007$ |
| AdaBoost | $0.586 \pm 0.009$ | $0.583 \pm 0.008$ | $0.603 \pm 0.007$ |
| kNN + RF | $0.594 \pm 0.008$ | $0.590 \pm 0.006$ | $0.597 \pm 0.008$ |
| FFNN | $0.604 \pm 0.010$ | $0.599 \pm 0.012$ | $0.611 \pm 0.011$ |
| DeepEnroll | $0.600 \pm 0.010$ | $0.598 \pm 0.007$ | $0.625 \pm 0.008$ |
| COMPOSE | $0.604 \pm 0.007$ | $0.597 \pm 0.006$ | $0.628 \pm 0.009$ |
| HINT | $0.629 \pm 0.008$ | $0.620 \pm 0.008$ | $0.645 \pm 0.006$ |
| SPOT | $0.685 \pm 0.010$ | $0.656 \pm 0.009$ | $0.630 \pm 0.007$ |
| HGGT | $0.6628 \pm 0.0158$ | $0.6880 \pm 0.0110$ | $0.6299 \pm 0.0088$ |
| HGT_aug | $0.7005 \pm 0.0152$ | $0.7057 \pm 0.0104$ | $0.6657 \pm 0.0102$ |
| HGGT_aug | $0.7114 \pm 0.0164$ | $0.7155 \pm 0.0107$ | $0.6787 \pm 0.0104$ |

**Table 3:**
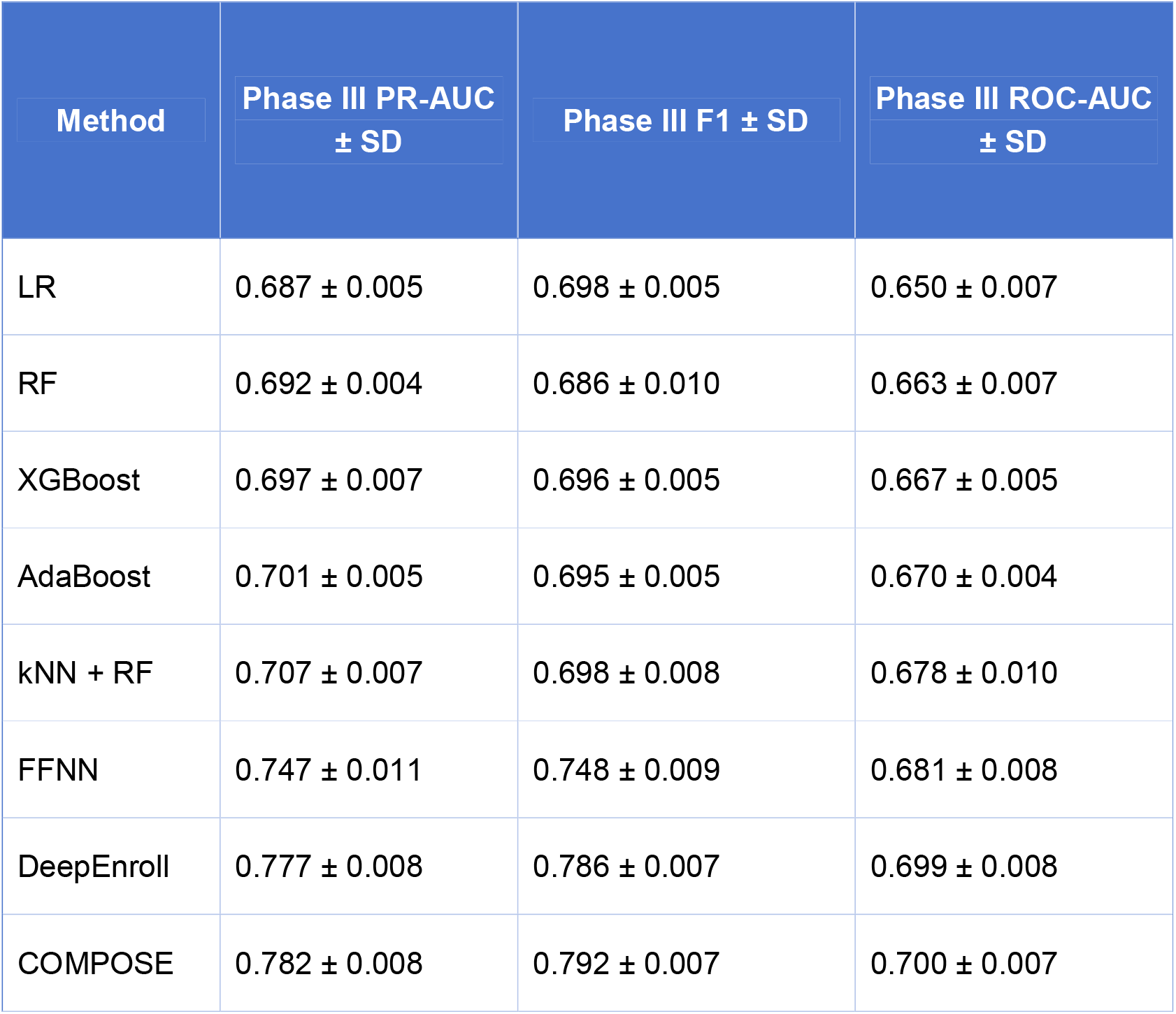
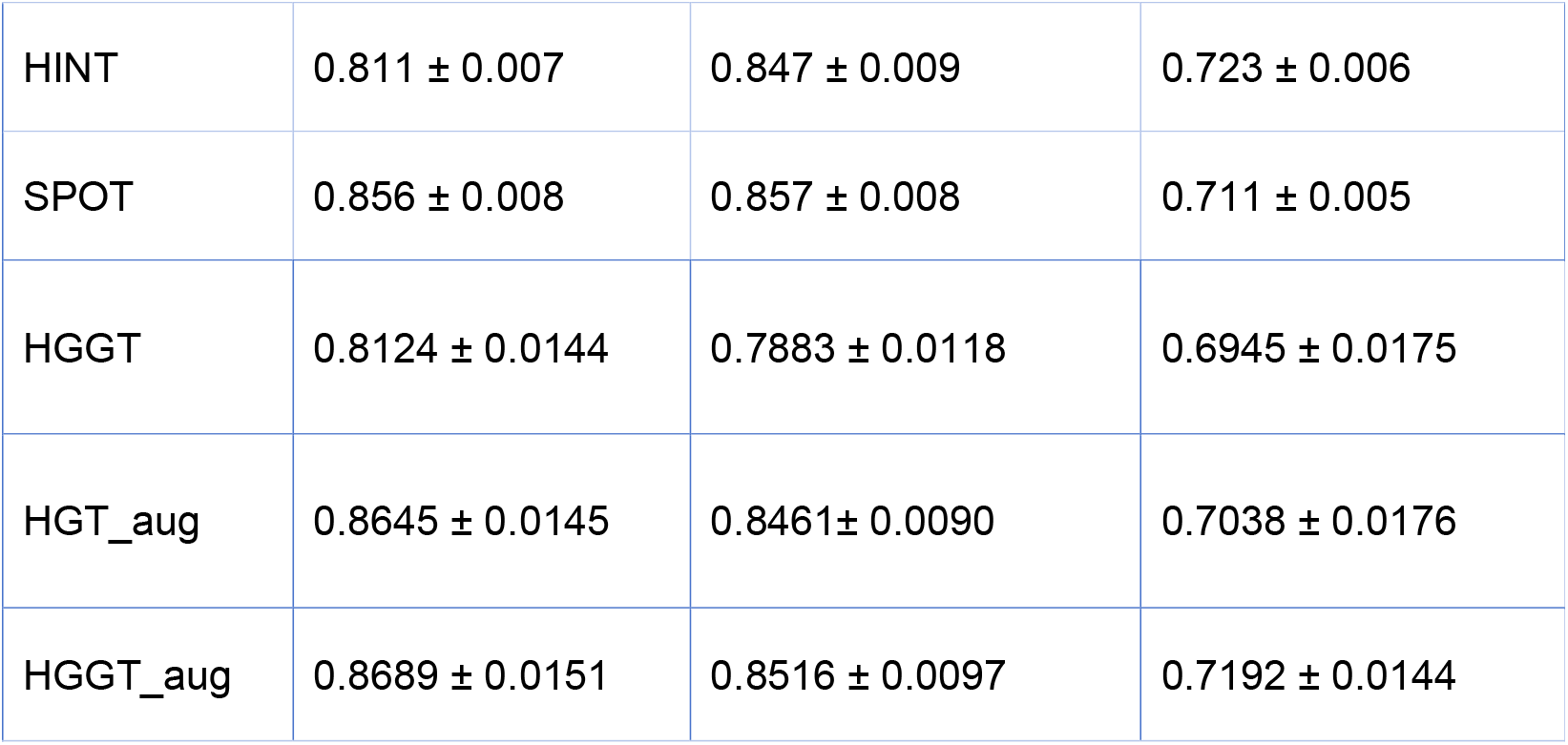
Performance comparison of different models on Phase III trials.

| Method | Phase III PR-AUC<br>± SD | Phase III F1 ± SD | Phase III ROC-AUC<br>± SD |
| --- | --- | --- | --- |
| LR | $0.687 \pm 0.005$ | $0.698 \pm 0.005$ | $0.650 \pm 0.007$ |
| RF | $0.692 \pm 0.004$ | $0.686 \pm 0.010$ | $0.663 \pm 0.007$ |
| XGBoost | $0.697 \pm 0.007$ | $0.696 \pm 0.005$ | $0.667 \pm 0.005$ |
| AdaBoost | $0.701 \pm 0.005$ | $0.695 \pm 0.005$ | $0.670 \pm 0.004$ |
| kNN + RF | $0.707 \pm 0.007$ | $0.698 \pm 0.008$ | $0.678 \pm 0.010$ |
| FFNN | $0.747 \pm 0.011$ | $0.748 \pm 0.009$ | $0.681 \pm 0.008$ |
| DeepEnroll | $0.777 \pm 0.008$ | $0.786 \pm 0.007$ | $0.699 \pm 0.008$ |
| COMPOSE | $0.782 \pm 0.008$ | $0.792 \pm 0.007$ | $0.700 \pm 0.007$ |
| HINT | $0.811 \pm 0.007$ | $0.847 \pm 0.009$ | $0.723 \pm 0.006$ |
| SPOT | $0.856 \pm 0.008$ | $0.857 \pm 0.008$ | $0.711 \pm 0.005$ |
| HGGT | $0.8124 \pm 0.0144$ | $0.7883 \pm 0.0118$ | $0.6945 \pm 0.0175$ |
| HGT_aug | $0.8645 \pm 0.0145$ | $0.8461 \pm 0.0090$ | $0.7038 \pm 0.0176$ |
| HGGT_aug | $0.8689 \pm 0.0151$ | $0.8516 \pm 0.0097$ | $0.7192 \pm 0.0144$ |

Three key metrics for evaluating predictive models are defined as follows: The Precision-Recall Area Under the Curve (PR-AUC) is derived from the Precision-Recall (PR) curve, which summarizes the trade-off between the true positive rate and the positive predictive value of a predictive model across different probability thresholds. The F1 score serves as the harmonic mean of precision and recall, offering a balanced measure that combines these two complementary metrics. Meanwhile, the Receiver Operating Characteristic Area Under the Curve (ROC-AUC) is based on the ROC curve, which captures the trade-off between the true-positive rate and the false-positive rate of a predictive model when using various probability thresholds.

Overall, across all three trial phases, the proposed HGGT architecture consistently outperforms the original HGT variant. Equipping HGGT with gene-level information as HGGT_aug, which embeds type-specific projection blocks and a trial-aware attention aggregation mechanism, yields the strongest performance on the TOP dataset. As shown in Table 1, HGGT already outperforms most conventional baselines in terms of PR-AUC and F1 and is competitive with the strongest non-graph method SPOT. Incorporating gene information consistently improves performance. HGT_aug improves over HGGT across all three metrics, and HGGT_aug further increases PR-AUC and F1 with a higher ROC-AUC than HGT_aug. As presented in Table 2, the gains of our architecture become more pronounced in Phase II trials. HGGT achieves competitive PR-AUC, F1 and ROC-AUC values that already surpass most baselines. Adding gene information to the original HGT improves performance, but HGGT_aug consistently yields the best results among all models and notably outperforms both HGT_aug and the strongest non-graph baseline SPOT. In Phase III trials shown in Table 3, HGGT remains highly competitive with strong PR-AUC and F1 values and performs on par with strong neural baselines. Incorporating gene information further boosts performance for both architectures. HGT_aug improves over HGGT across all three metrics, and HGGT_aug attains the best overall PR-AUC and ROC-AUC and surpasses SPOT. These results demonstrate that both the heterogeneous graph design of HGGT and the integration of gene and target information provide substantial and robust gains for clinical trial outcome prediction.

## 6. Conclusion

The HGGT model’s performance demonstrates the value of heterogeneous graph representations in capturing the multifaceted determinants of clinical trial success. By integrating biological entities (genes, targets) with trial-specific information (abstracts, criteria), the model leverages both structured and unstructured data to make robust predictions. The model’s robust performance across metrics, particularly its high recall and phase-specific adaptability, highlights the potential of deep graph-based learning in optimizing clinical trial design. By capturing complex relationships between biological entities and trial features, the HGGT approach offers a promising tool to reduce the cost and time of drug development, accelerating access to new therapies. Limitations include reliance on public trial data, which may underrepresent failed trials due to reporting biases. Future work could incorporate additional data sources (e.g., real-world evidence) and explore explainability methods to identify key features driving predictions, thereby enhancing trust among clinicians and regulators.

## Data availability

The data used and/or analyzed during the current study are available from the corresponding author upon reasonable request.

